# Scenario projections of congenital syphilis incidence in California, 2025–2030

**DOI:** 10.64898/2026.04.30.26352168

**Authors:** Natalie M. Linton, Nicole O. Burghardt, Natalie S. de Guzman, Robert E. Snyder, Tomás M. León, Kathleen Jacobson

## Abstract

**Background:** In 2022, the yearly rate of congenital syphilis (CS) cases in California reached its highest point since 1950. We explored how different prevention strategies in California could impact future CS case incidence.

**Methods:** Scenario modeling was used to project CS incidence from 2025 through 2030 based on 2015–2024 surveillance data. We compared a baseline scenario with scenarios that modeled 1) changes in the rate of annual female syphilis cases; and 2) changes in the proportion of pregnant persons adequately treated for syphilis. We then used 2016–2023 hospitalization discharge data from California to estimate charges associated with treatment of a CS case.

**Results:** A 5–50% reduction in the absolute rate of female syphilis cases led to a 3–30% decrease in cumulative CS cases during the projection period. Increasing the proportion of pregnant persons adequately treated for syphilis by 5–50% led to a 2–23% decrease; combining both scenarios resulted in a 4–45% reduction. In the baseline scenario, projected CS treatment-associated hospitalization charges were estimated to total $399 million. A 45% reduction in CS cases resulting from the combined interventions would reduce charges by $158 million.

**Discussion:** Female syphilis incidence and adequate treatment during pregnancy have substantial impact on CS case incidence and associated charges. California has implemented universal syphilis screening guidelines for all people aged 15–49 years alongside interventions supporting testing and treatment of pregnant persons.

Continued implementation of these interventions will result in considerable savings around CS treatment and reduced need for management of long-term sequelae.

## Introduction

Most cases of congenital syphilis (CS)—vertical transmission of *Treponema pallidum*, the causative agent of syphilis—can be prevented with timely diagnosis and adequate treatment of pregnant persons.^1^ However, CS case rates in the United States increased precipitously between 2010–2022.^2^ In California, the yearly rate of CS increased by an average of 33% each year between 2013 and 2022, from 11.7 cases per 100,000 live births in 2013 to a peak of 147.7 cases per 100,000 live births in 2022, reaching its highest point since 1950.^3^ CS can cause a myriad of symptoms in utero, leading to stillbirth and fetal loss or—for children who survive and are born with CS—bone abnormalities, organ enlargement, blindness, hearing loss, and other potential birth defects.

To combat rising cases, health departments across California focused on implementing and scaling up interventions to reduce the number of acquired syphilis cases and improve treatment for females infected during pregnancy. Co-occurring circumstances that may have affected incidence and treatment completion include delayed healthcare seeking during the coronavirus disease 2019 (COVID-19) pandemic, use of doxycycline for treatment of non-pregnant persons during the shortage of benzathine penicillin G in 2023– 2024, and use of doxycycline for non-syphilis related conditions (which may have inadvertently treated undiagnosed syphilis cases). In recognition of the severity of the situation nationally, the U.S. Department of Health and Human Services established a National Syphilis and Congenital Syphilis Syndemic Federal Task Force.^4,5^

Then, in 2023, CS rates began to decrease in California and seventeen other states,^6^ providing hope that interventions were beginning to work. Interventions and other circumstances that may have contributed to the 18% decline in CS cases and 53% decrease in primary and secondary acquired male and female syphilis cases in California between 2023–2024 are shown in Table 1. In one notable example, the City and County of San Francisco saw a 51.4% decline in early syphilis cases among men who have sex with men and transgender women during the year following the implementation of doxycycline postexposure prophylaxis (DoxyPEP) guidelines.^7^ A similar reduction (53.1%) in cisgender male syphilis cases reported in Seattle is believed to have indirectly decreased transmission to cisgender females from shared sexual networks between cisgender women and people on doxy-PEP ^8–10^

**Table 1.** Interventions and circumstances in California with the potential to affect congenital syphilis case incidence, 2019–2024.

| Interventions and circumstances | Cases | Treatment |
| --- | --- | --- |
| First statewide DoxyPEP recommendations to administer DoxyPEP to prevent and syphilis | ✓ |  |
| Doxycycline administration for other conditions (e.g., chlamydia, pneumonia) indirectly working as DoxyPEP for syphilis | ✓ |  |
| Benzathine penicillin G shortage (2023–2024) led to more non-pregnant people receiving treatment with oral doxycycline, which requires fewer healthcare interactions compared to Benzathine penicillin G for treatment of late syphilis | ✓ |  |
| Delayed healthcare seeking activities during the COVID-19 pandemic | ✓ | ✓ |
| Increased funding to develop, train, and support healthcare providers and STI programs in local health departments enabled increased and enhanced patient and partner services | ✓ | ✓ |
| Sexually transmitted infection screening and treatment offered in non-traditional settings (e.g., in emergency departments, jails, and homeless encampments) | ✓ | ✓ |
| Increased access to prenatal care, including prenatal care services in non-traditional settings (e.g., in emergency departments, jails, and homeless encampments) | ✓ | ✓ |
| Individualized outreach to test and treat pregnant and non-pregnant persons in non-traditional settings<sup>32</sup> (e.g., in fast food restaurant bathrooms, gas stations, the backseats of cars under bridges, etc.) | ✓ | ✓ |

Cases: The intervention is expected to affect female case incidence.
Treatment: The intervention is expected to affect adequate treatment in pregnancy. Many of the interventions that would impact treatment adequacy were introduced in 2020 or later.
COVID-19: coronavirus disease 2019.
DoxyPEP: pre- or post-exposure doxycycline prophylaxis.

Despite progress towards reducing syphilis rates in California, numerous factors continue to contribute to the late initiation of prenatal care in pregnant people or inhibit their timely testing and adequate treatment, such as methamphetamine and injection drug use, incarceration, and homelessness.^11,12^ When birthing parents are not adequately treated for syphilis during pregnancy, infants are recommended to be hospitalized to receive CS treatment.^13^ Excess cost per hospitalized CS case among newborns in the United States was estimated at $14,576 in 2024 dollars during 2005–2009 and rose to an average of $64,345 in 2024 dollars for hospitalizations between 2009–2016.^14–16^ Another newborn hospitalization study in the United States between 2016–2020 found hospitalization costs for newborns with CS were 4.9 times the cost of newborn hospitalizations without CS.^17^ Compared to hospitalizations without a CS diagnosis, hospitalizations with a CS diagnosis had a greater odds of being uninsured or claimed to public insurance, and greater odds of a household income of less than $25,000 per year.^16^

To help assess the impact that interventions with varying success rates might have on future CS incidence and prioritize the most impactful CS prevention activities, we simulated future projections of CS cases based on intervention effect using a scenario modeling framework.^18^ The framework explores the individual and combined effects of two types of interventions that would be implemented in California to prevent CS during 2025–2030: 1) changes in the rate of annual female syphilis cases; and 2) changes in the proportion of pregnant persons adequately treated for syphilis. We also obtained data on hospitalization charges likely to represent treatment of CS in California between 2016–2023 and inferred a range of potential future charges associated with CS treatment based on this information.

## Methods

Acquired and congenital syphilis surveillance case data follow Council of State and Territorial Epidemiologist case definitions. For acquired syphilis cases, the four stages of infection are described in greater detail in the Supplementary Materials. We simplified these stages into early syphilis (primary, secondary, and early non-primary non-secondary syphilis) and unknown duration or late syphilis (UDLS). Early syphilis cases require evidence that the infection was acquired in the last twelve months.

Syphilis stage was incorporated into the modeling framework because treatment protocols during pregnancy differ by disease stage. Pregnant individuals with early syphilis need only receive a single dose of benzathine penicillin G to be considered adequately treated for syphilis, while pregnant individuals diagnosed with UDLS must receive three doses of benzathine penicillin G spaced one week apart.^19–21^ The risk of vertical syphilis transmission also differs by acquired syphilis stage, with risk highest in the early stages of infection and declining over time since infection.^22^

### Model and scenarios

Our scenario modeling framework (Figure 1) was developed to project CS case incidence in California using state-level public health surveillance data from 2015–2024 based on user-defined assumptions of changes to future female case rates and treatment adequacy in pregnant persons The model presented in this study does not capture transmission dynamics of acquired syphilis, and assumptions used when deciding on scenario parameters reflected subject matter expert opinion of the potential range of impact of current and future interventions during the projection years.

**Figure 1.**
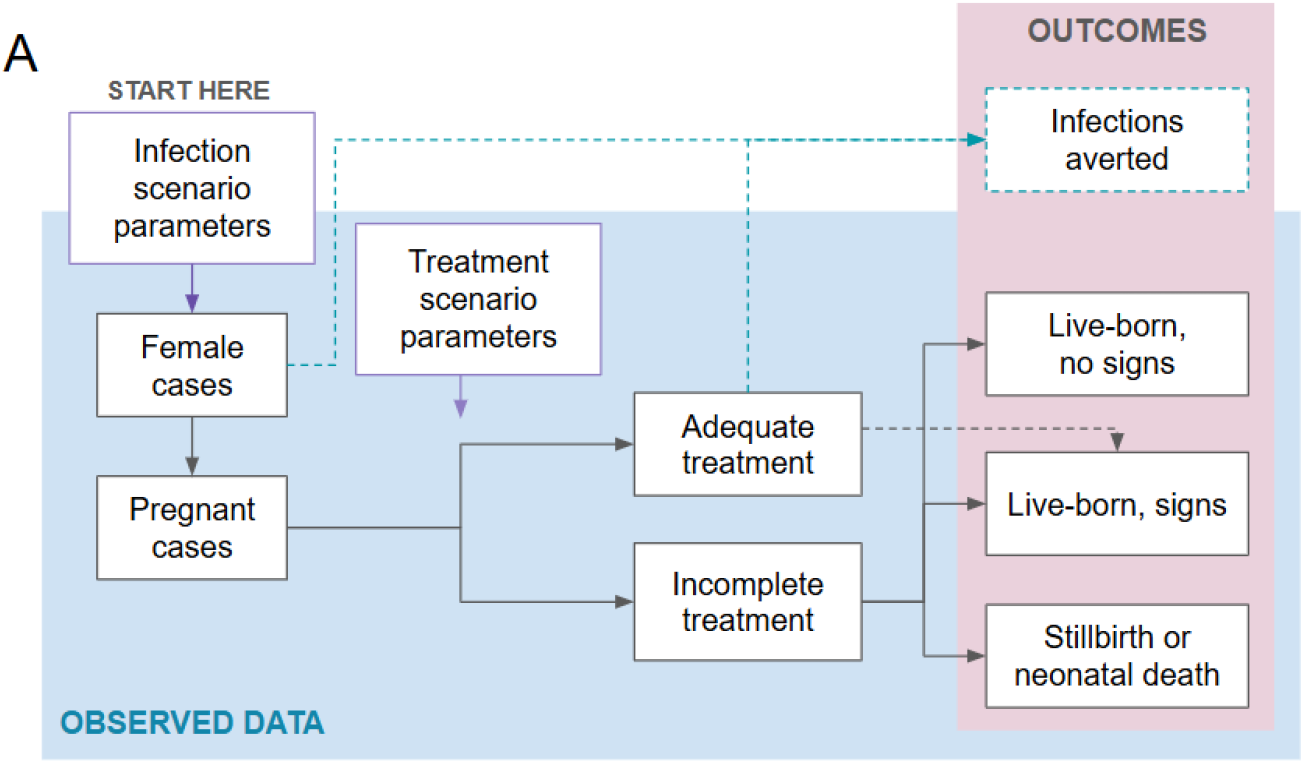
Workflow of scenario projection model framework, starting with an assumption about the number of female cases and following through to congenital syphilis outcomes. Solid lines represent transitions quantified directly from surveillance data. Dotted lines represent transitions that were not quantified from surveillance data that were calculated within the model.

### Parameters

The model includes two types of scenario parameters: one that influences projections of female acquired syphilis cases, and another that affects the proportion of pregnant cases who receive adequate treatment for syphilis during pregnancy (as defined in the Supplementary Materials). The model takes as input a percentage change (increase, decrease, or no change) in female case rates and the proportion of pregnant cases adequately treated during pregnancy. These changes were app recent reported data point (2024 in this study) and were specified to be applied annually or across the entire projection period. Table 2 illustrates this scenario structure.

**Table 2.**
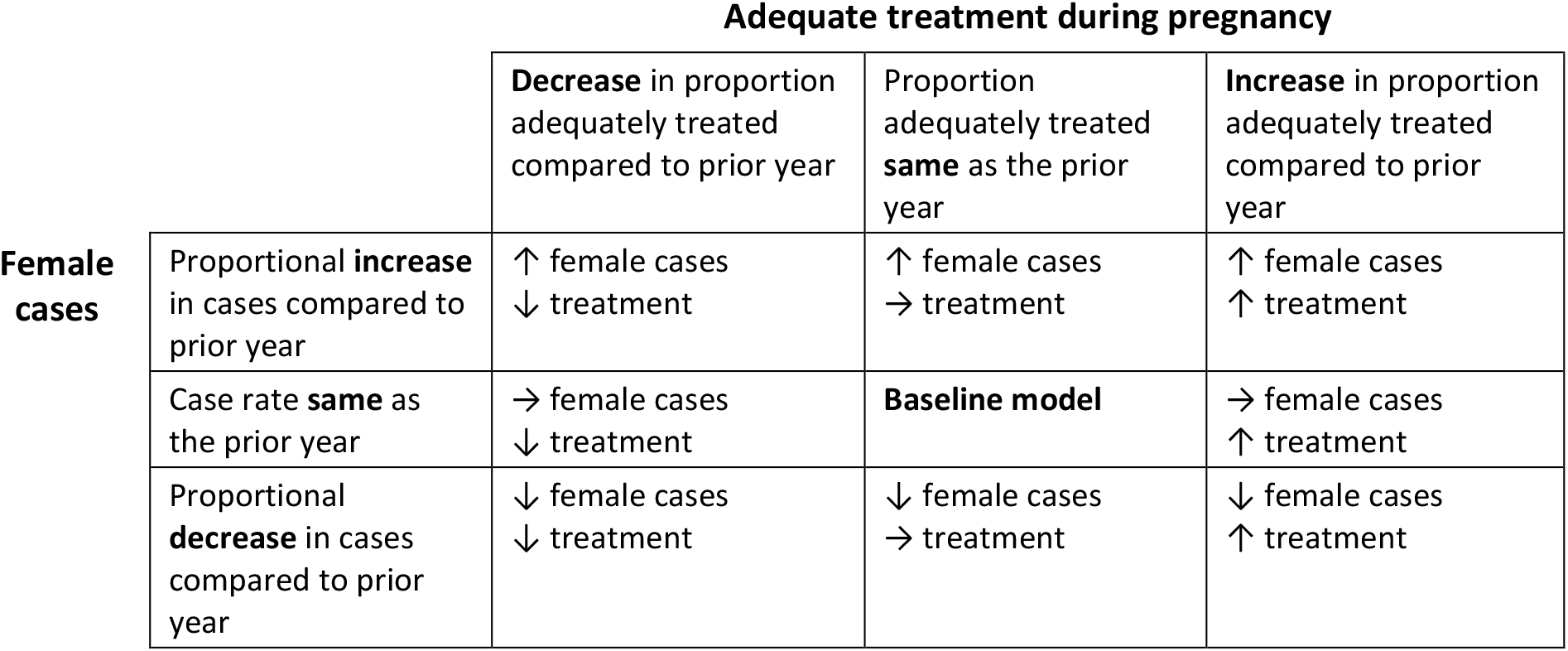
Overview of the scenario structure.

The remaining parameters used to derive the number of future CS cases and timing of their diagnoses are described in detail in the Supplementary Materials. Projections were repeated across 1000 simulations per scenario and we present the median outcome across all simulations for a given scenario as well as the 80% projection interval, which reflects the spread in the distribution of outcomes across the projected trajectories.

### Hospitalization data

Hospitalization discharge summary data for 2016–2023 were derived from the California Department of Health Care Access and Information Patient Discharge Dataset (PDD). The PDD includes records of all inpatient discharges from California-licensed hospitals, including general acute care, acute psychiatric, chemical dependency recovery, and psychiatric health facilities. It does not include Veteran’s Administration hospitals. CS hospitalizations were defined as having ICD-10-CM code A.50.* in any of 25 diagnosis fields. Hospitalization charges were adjusted to 2024 US dollars using the priceR package.^15^

Temporally, we evaluated only admissions that occurred within 28 days of birth. There is no best practice for obtaining charges related to the recommended 10–14 days of hospitalization after birth for infants diagnosed with CS to receive treatment. Depending on the hospital’s testing practices, delays in the receipt of test results for the birthing parent or infant could potentially delay starting treatment. However, not all admissions occurred within the first day of birth and some admissions are considered transfers where the initial hospitalization had a primary diagnosis of birth and the secondary hospitalization had a primary diagnosis of CS as well as procedure codes and length of stay (10–14 days) consistent with CS treatment. We use this subset of hospitalizations to represent an estimate of hospitalization charges representing treatment for CS.

### Data availability

The case data used in this study are not publicly available. However, parameters fitted to these data and code that will allow for reproduction of the study using publicly available Centers for Disease Control and Prevention AtlasPlus data are available at https://github.com/cdphmodeling/congenital_syphilis_projection_model.

## Results

Figure 2A shows the number of acquired syphilis cases per year among all individuals assigned female at birth of childbearing age (ages 15–44) in California. Case rates were higher for ULDS compared to early syphilis (Figure 2B) and increased proportionally from 2015 through 2021. In 2022, the relative rate of increase since 2015 for early syphilis began to decrease while the rate of UDLS continued to increase through 2023 before plateauing between 2023 and 2024 (Figure 2C). The exception was 2020, when the rate of syphilis decreased due to reduced syphilis screening and subsequent diagnoses in the first year of the COVID-19 pandemic.^23^

**Figure 2.**
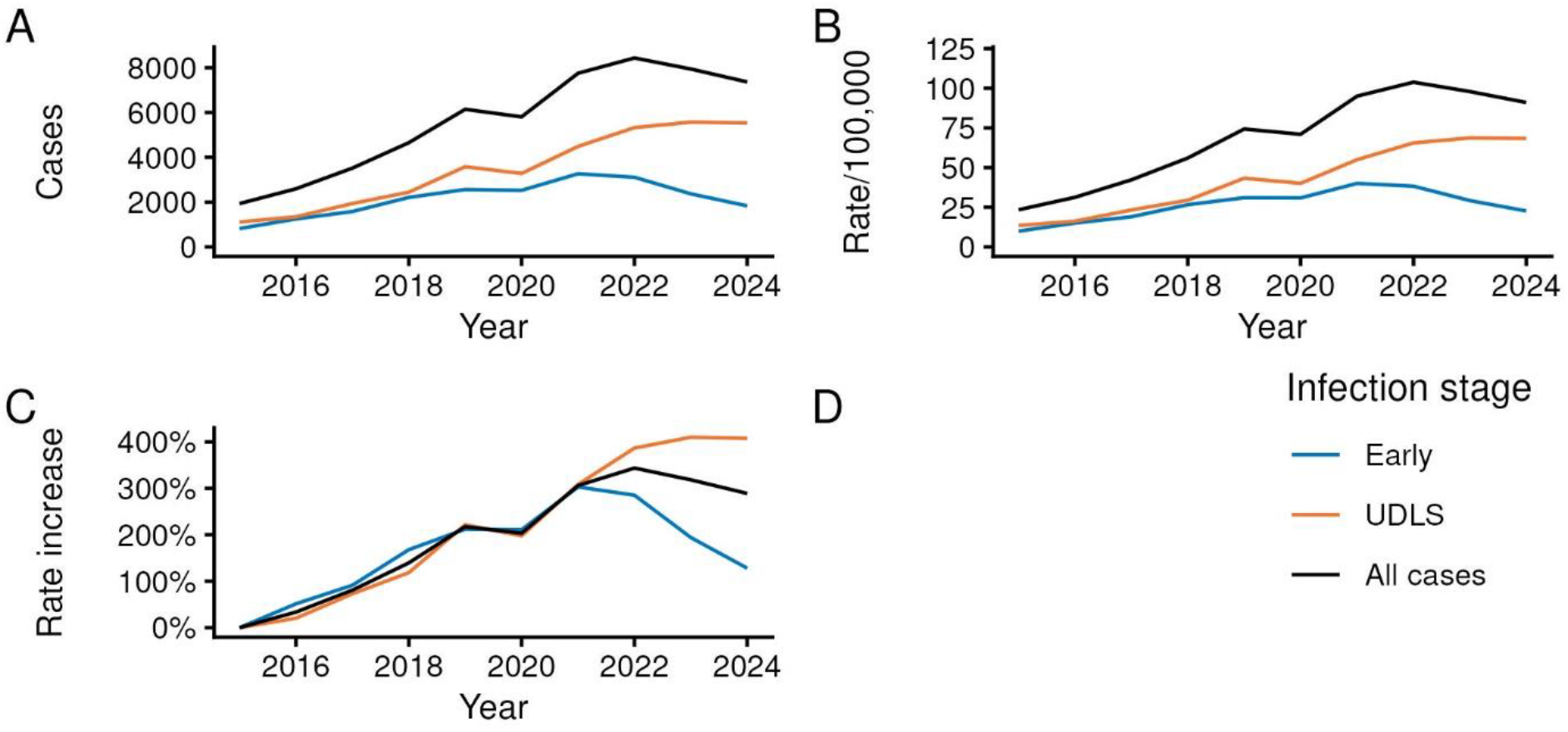
Yearly rate of acquired syphilis by stage—early syphilis and unknown duration/late syphilis (UDLS)—for females 15–44 years old in California, 2015–2024

Under the baseline scenario, we estimated a median cumulative total number of CS cases between 2025– 2030 of 3,009 (80% projection interval [PI]: 2,736–3,293, averaging 502 cases per year) across all simulations (Table 3). A best-case scenario with a 50% reduction in female cases and 50% increase in treatment adequacy among pregnant persons reduced the cumulative projected cases by 45%, to 1,667 (80% PI: 1,511–1,856), or an average of 278 cases per year. Keeping the 50% reduction in female cases with no change in the proportion of pregnant persons adequately treated since 2024 limited the reduction in cumulative cases from the baseline model to a 30% decrease, or 2,118 (80% PI: 1,912–2,319) cases over the projection period.

**Table 3.**
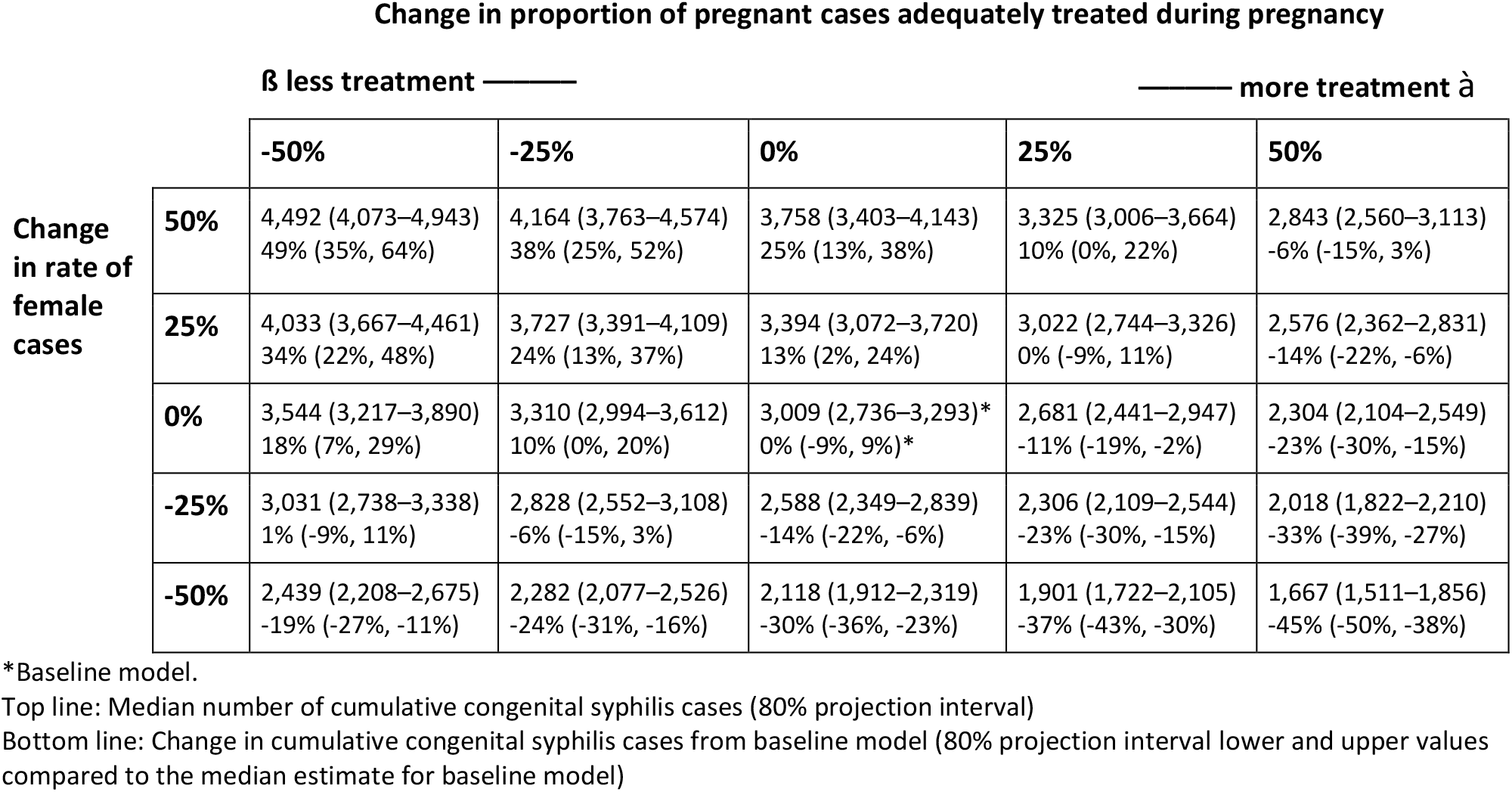
Estimates of cumulative congenital syphilis cases and change in cumulative cases compared to baseline model median from 2025–2030 in California.

A 50% increase in female cases and 50% decrease in treatment adequacy among pregnant persons between 2025–2030 (i.e. the worst-case scenario presented here) increased the projected number of CS cases by 49% to a cumulative 4,492 (80% PI: 4,073–4,943) cases, or an average of 749 cases per year.

Maintaining the proportion of pregnant persons who were adequately treated constant reduced the increase in cumulative cases from 49% to 25%, or 3,758 (80% PI: 3,403–4,143) cases between 2025–2030. A 50% increase in the proportion adequately treated offset the increase in female cases and led to a net 6% reduction in cumulative CS cases.

Comparing the relative effects of the two scenario parameters, a proportion change in female cases that would reflect interventions losing effectiveness (i.e., positive parameter value) led to more cumulative CS cases over the projection period compared to an equivalent loss of intervention effectiveness for the adequate treatment scenario parameters (i.e. negative parameter value). For example, a 50% increase in female cases with no change in the proportion adequately treated since 2024 led to a 25% increase in cumulative CS cases while a 50% reduction in the proportion adequately treated during pregnancy and no change in female cases since 2024 only led to a 18% increase in cumulative CS cases between 2025–2030.

The relative effect of the two scenario parameters when the parameters reflected interventions increasing in effectiveness over time (for the female cases parameter, a value < 0, and for the treatment adequacy parameter, a value > 0) similarly saw better outcomes for interventions targeting female acquired syphilis incidence. For example, a 50% decrease in female cases with no change in the proportion of pregnant persons adequately treated since 2024 led to a 30% decrease in cumulative CS cases, while no change in female cases but a 50% increase in adequate treatment led to a 23% decrease in cumulative CS cases.

Figure 3 shows the trajectory of median CS cases reported each year between 2025–2030 under the various scenarios with the same ordinal structure as Table 2 and Table 3. The center figure in grey is the baseline scenario where both scenario parameters (female cases and proportion of pregnant cases adequately treated during pregnancy) are kept constant at their 2024 values. The upper left figure contains median trajectories and 80% PI shading for all scenarios with an increase in female cases and decrease in proportion of cases adequately treated during pregnancy. Similarly, the figure on the bottom right contains median trajectories and 80% PI shading for all scenarios with a decrease in female cases and increase in the proportion of cases adequately treated during pregnancy.

**Figure 3.**
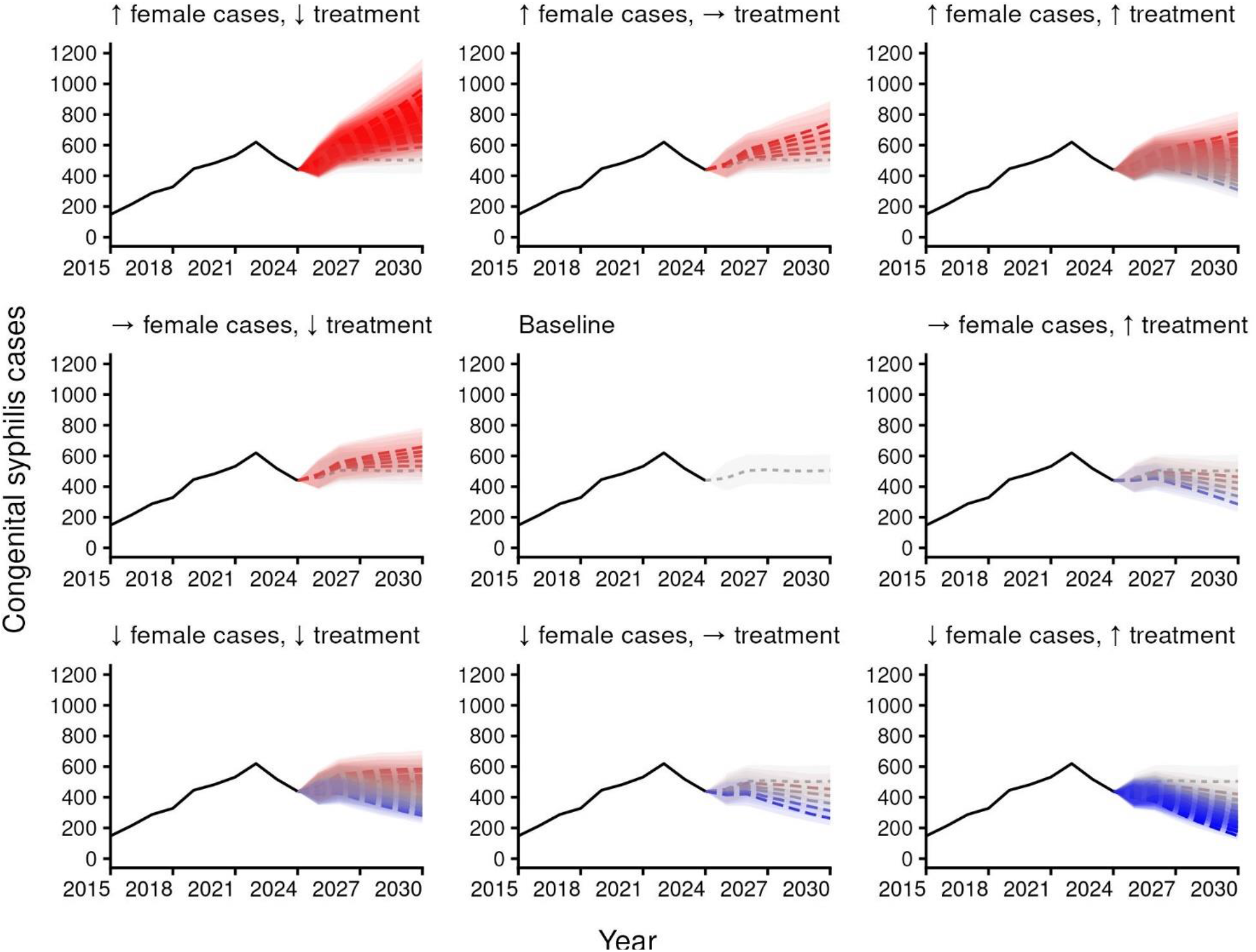
Median trajectories and 80% projection intervals of yearly CS cases in California from 2025–2030 by scenario type. A down arrow (↓) indicates the parameter has a 5–50% decrease and an up arrow (↑) indicates a 5–50% increase in the parameter value over the projection period from the last reported value in 2024. A right arrow (→) indicates the 2024 parameter value is used for all projection years. Reported case numbers are in black. Darker red or blue color indicates greater increase (red) or decrease (blue) in cumulative CS cases over the projection period compared to the baseline model. Trajectories with colors closer to grey have cumulative case estimates similar to the baseline model.

Reflecting the results in Table 3, the scenario trajectories depicted in Figure 3 illustrate how decreases in intervention effectiveness targeting prevention of infections among females in California (top row) can reverse the decreases in CS cases seen in 2023 and 2024, and potentially re-initiate the rapid increase in cases seen between 2015–2022. Scenarios where effectiveness of both types of intervention continues to improve, further reducing female cases and increasing adequate treatment for pregnant cases (bottom right) continue the decrease in CS cases that began in 2023.

Figure 4 shows the total change in cumulative CS cases compared to the baseline scenario along each intervention axis. The baseline scenario (0% change in female cases and 0% change in adequate treatment for pregnant persons) is shown in the center, and scenarios range along a 50% additional reduction/increase in female cases and 50% reduction/increase in adequate treatment for pregnant persons between 2025–2030.

**Figure 4.**
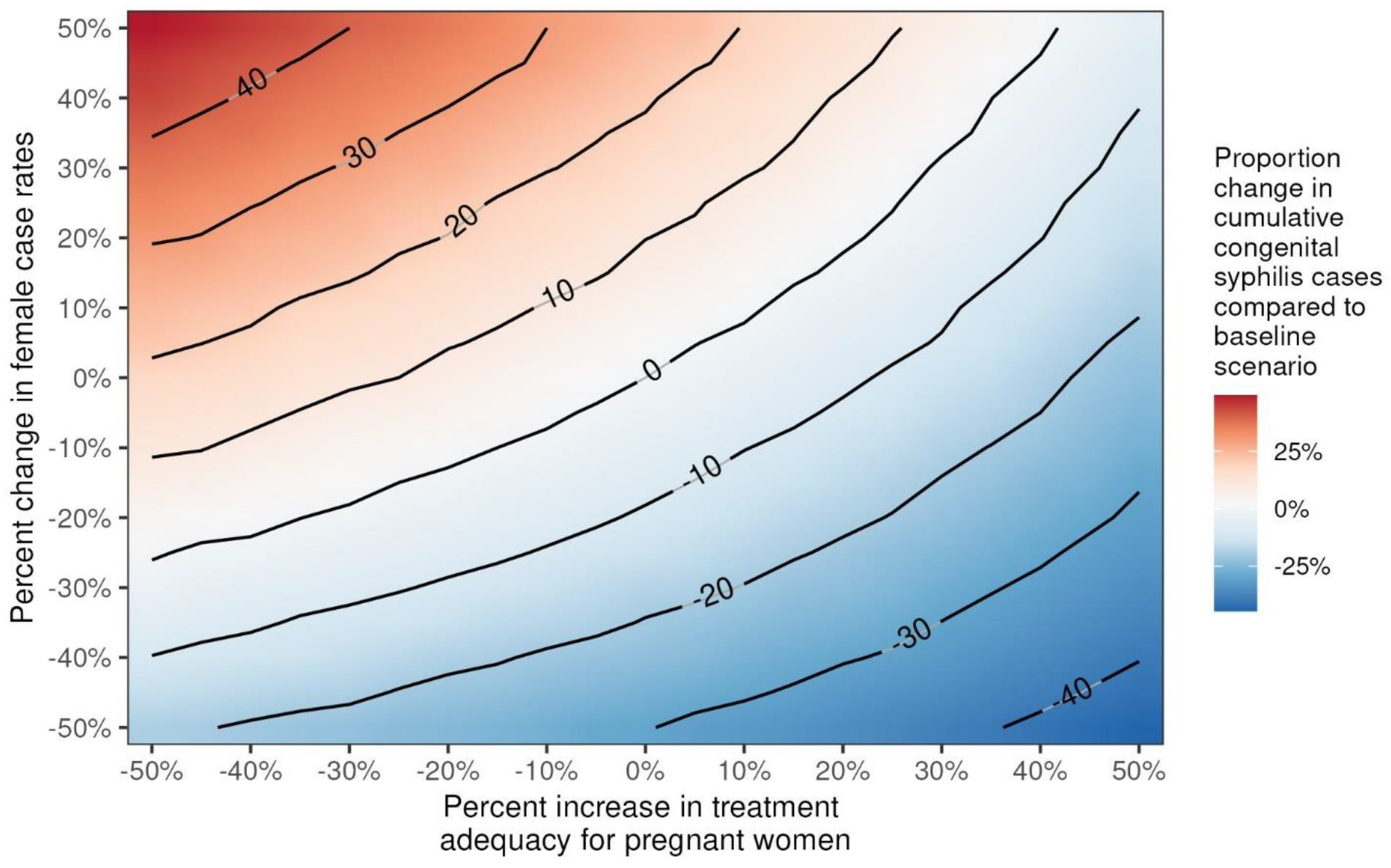
Percent reduction in cumulative projected congenital syphilis (CS) cases between 2025–2030 by scale of interventions affecting female syphilis case rates and treatment adequacy for pregnant persons. Contour lines are shown for every 10% difference in cumulative CS cases.

### Hospitalization charges and projected charges

Between 2016–2023, the total charges for the 4,667 CS hospitalizations identified in California’s hospitalization discharge dataset totaled $1.15 billion in 2024 dollars. We found more hospitalizations with CS diagnoses than there were reported CS cases, even after attempting to deduplicate hospitalizations to the infant level (Figure S8). The mean charge per hospitalization was $260,830 (95% confidence interval [CI]: $232,617–$260,876). Length of stay per hospitalization ranged from 0 to 369 days. Most charges (71%) over the seven-year period were to Medi-Cal, and 22% were to other government payers, which included any non-Medi-Cal form of payment from government agencies, such as the California Children’s Services or Tricare.

We obtained a subset (n=34) of hospitalizations where the infant had an initial hospitalization with a primary diagnosis associated with birth and a second hospitalization with a primary diagnosis of CS and 10– 14-day length of stay initiated within the first 28 days of birth. These hospitalizations were likely for CS treatment. The average charge for those hospitalizations was $144,421 (95% CI: $124,124–$164,718) and most charges (74%) were to Medi-Cal.

In the baseline scenario, hospitalization charges for projected CS cases that would require treatment for CS (i.e. live-born cases, excluding stillbirths) were estimated to total approximately $399 million in 2024 dollars between 2025–2030 for California. A 30% reduction in live-born CS cases based on a 50% decrease in female cases and no change to the proportion of pregnant cases adequately treated for syphilis during pregnancy would reduce these charges by $118 million to $281 million over the six-year projection period. A 23% reduction in cumulative live-birth CS cases based on a 50% increase in the proportion of pregnant cases adequately treated for syphilis during pregnancy and no change to reported female cases from 2024 values would reduce estimated treatment charges by $66 million to $333 million. A best-case scenario 45% reduction in cumulative projected CS cases based on a 50% decrease in female cases and 50% increase in the proportion of pregnant cases adequately treated during pregnancy would reduce CS treatment charges by $158 million to $241 million. In contrast, a worst-case scenario 49% increase in cumulative CS cases based on a 50% increase in female cases and 50% decrease in the proportion of pregnant cases adequately treated during pregnancy would increase projected treatment-associated charges by $250 million to $649 million between 2025–2030.

## Discussion

Using scenario modeling we demonstrate that reducing the rate of syphilis among females and increasing treatment adequacy for pregnant persons are both impactful interventions to reduce future CS incidence, though the former was projected to result in a greater overall decrease in future CS cases. Reducing incidence among female cases and increasing adequate treatment among pregnant persons by 50% each between 2025 and 2030 could prevent 45% of CS cases that would otherwise occur were current 2024 estimates to be held static. Prevention of these cases could reduce hospitalization charges associated with a CS diagnosis by an estimated $331 million between 2025 and 2030.

For this analysis we did not model a specific intervention. Rather, the model propagated user-specified assumptions regarding an increase or decrease in intervention effectiveness over the projection period to quantify the downstream effect on CS case incidence. Examples of recently introduced interventions are described in Table 1, though longstanding interventions such as screening for syphilis in the first trimester of pregnancy at the first encounter as indicated in the California Department of Public Health syphilis screening guidelines are also critical to include in selecting scenario parameters.^24^ The modeling framework could be expanded further to allow for non-linear user-defined assumptions of intervention effects^25^ or by utilizing forecasting methods.^26,27^ A more complex modeling framework could include a transmission dynamics model that outputs yearly cases in females ages 15–44 as an input for the rest of the CS modeling framework. Alternatively, rather than estimating future female cases and determining the number of pregnant persons among them, the model could be adapted to estimate the number of pregnant persons and then estimate syphilis prevalence.^28^

From 2022–2024, the rate of female early syphilis cases decreased while the rate of UDLS cases continued to increase to a plateau in California. The difference in trends between these two groups may be due to increases in the rate of syphilis screening that has led to the detection of more cases with unknown or long-term infection who would otherwise have gone undetected. The wide-ranging list of interventions listed in Table 1 suggest that syphilis (and therefore CS) cases could continue to decrease in California (Figure S3).

CS is largely preventable with timely diagnosis and treatment of the birthing parent during pregnancy,^1,29^ but even with timely and adequate treatment of the birthing parent, 8–12% of infants still meet the CS surveillance case definition via the infant criteria alone.^30^ As such, upstream prevention of syphilis infection among females is a better opportunity to prevent CS than adequately treating pregnant persons already infected with syphilis and our modeling supports there is a greater downstream impact in preventing CS cases when interventions focus on preventing cases in females. The effort/workload required to achieve success in these interventions is difficult to assess as they are interrelated and impacted by a host of factors including but not limited to comprehensive partner services to prevent ongoing transmission, engagement in early prenatal care, and the resources for local public health teams to meet populations where they are at out in the community.

Hospitalization charges associated with a CS diagnosis have increased over recent decades.^14,16 17^ We estimated that in California each live-birth case of CS prevented would avoid an average of $144,421 in hospitalization charges for CS treatment. This value only estimates immediate treatment costs and not longer-term medical, educational, and other specialized care that may result from CS sequelae.^17^ These costs could be represented as quality adjusted life years (QALYs), which measures the quality and the quantity of life lived as a measure of disease burden. QALY loss is minimal for infants that survive infection but are more costly when infants survive but have life-long, costly disabilities that yield significant costs to the individual and medical systems, or are born alive but die in the days or weeks following birth, or die before birth.^31^ QALYs were not quantified in this report, but could be estimated based on model results (see Supplementary Materials). Preventing transmission of syphilis to females, by reducing the incidence in both males and females, would dramatically reduce societal and economic costs due to CS.

Our model shows that the most important upstream intervention to reducing CS is to reduce the rate of syphilis among females. We demonstrate that with the maximal combined interventions preventing 45% of CS cases would consequently save $241 million in CS treatment-associated hospital charges from 2025 through 2030. To this end, syphilis screening and treatment must be enhanced, beginning with the implementation of guidance recommended by the California Department of Public Health to screen all people aged 15–44 at least once in their lifetime and then offer screening annually thereafter.^32^

## Data Availability

Publicly available data are at https://gis.cdc.gov/grasp/nchhstpatlas/tables.html
Parameters that can be used to reproduce the study in combination with publicly available versions of the data are at https://github.com/cdphmodeling/congenital_syphilis_projection_model

