## Supplement for "Scenario projections of congenital syphilis incidence in California, 2025–2030"

**Supplementary Materials**

This document provides the supplementary materials for the study "Scenario projections of congenital syphilis in California, 2025–2030." Code and data can be obtained from cdphmodeling/congenital_syphilis_projection_model

**Data and disease information**

In this analysis, female syphilis cases include all individuals assigned female at birth (hereafter, females), though in California congenital syphilis (CS) cases to date have only been born to female syphilis cases who identified as women.

Syphilis is diagnosed as one of four stages:

- Primary syphilis: One or more ulcerative lesions (e.g. chancre) and a positive laboratory test indicative of syphilis.
- Secondary syphilis: Symptoms and signs possibly indicating secondary syphilis and a positive laboratory test indicative of syphilis.
- Early non-primary non-secondary syphilis (ENPNS): infection within the 12 months preceding diagnosis, but no signs or symptoms of primary or secondary syphilis, and a positive laboratory test indicative of syphilis.
- Unknown duration or late syphilis (UDLS)**:** no clinical signs or symptoms of primary or secondary syphilis, with epidemiological and laboratory information indicating infection occurred >12 months previously, or there is insufficient evidence to conclude that infection was acquired during the 12 months preceding diagnosis.

Neurosyphilis is another stage but was not included due to rarity of the condition. Infection trends are similar across early syphilis stages (primary, secondary, and ENPNS) so these were combined stages to simplify the model for projecting infections among those assigned female at birth.

**Model workflow**

The following section describes parameters used in the model in the order that they are used in the model workflow (Figure 1A as shown in the main text). Once total female cases are projected for each of the years 2025–2030, we use the *MCMCprecision* package to fit a Dirichlet distribution to 2015–2024 proportions of female syphilis cases of childbearing age (15–44) by stage at diagnosis (early or who were pregnant while infected (Figure S1A and B). In California, the options for this variable are Y (yes, pregnant), N (no, not pregnant), and U (unknown if pregnant). Cases reported to have unknown pregnancy status were considered to not be pregnant and recategorized as not pregnant. Although it would be possible to use a beta distribution for fitting, we retain the use of a Dirichlet distribution so that the option to use >2 categories for fitting remains open.

We sample from the Dirichlet distribution to obtain proportions of females diagnosed with syphilis who are pregnant at the time of their diagnosis, and as shown in Figure S1C the median value of the samples across all simulations are fairly consistent across projection years, however each individual sample will affect results for subsequent parameters being sampled, as described in the remaining paragraphs on methods.

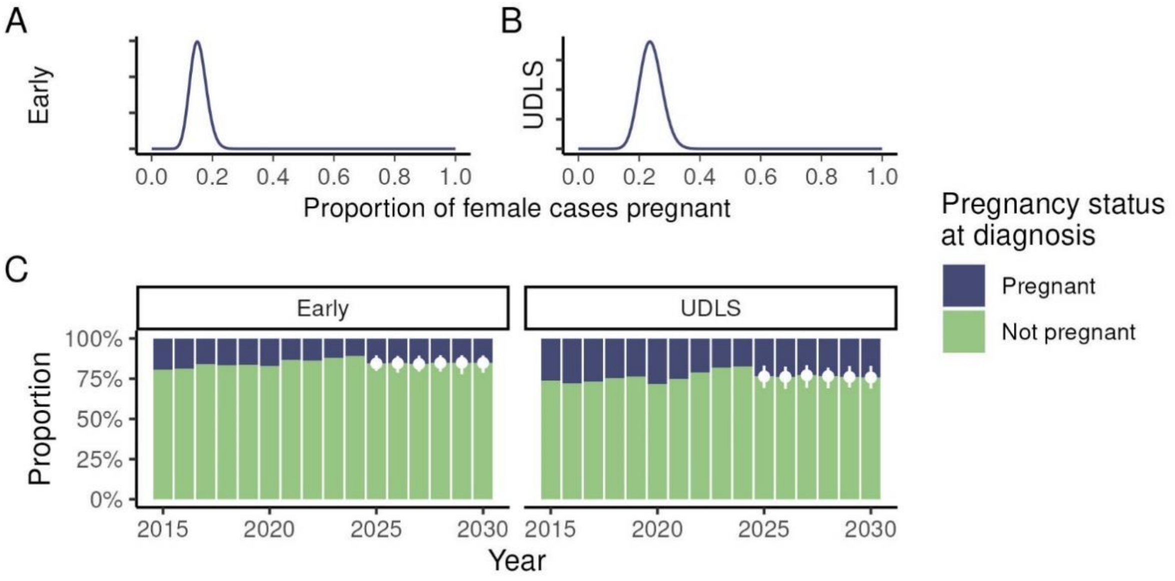


Figure S1. Reported (2015–2024) and projected (2025–2030) distributions (A, B) and proportions (C) of female acquired syphilis cases pregnant by stage of infection at diagnosis

After the number of pregnant individuals in each projection year is determined, each of these pregnant cases is assigned a date of diagnosis that is multinomially distribution with equal probability for each day of their diagnosis year. We chose to use equal probability for each because looking at our historical data there was no clear temporal pattern from month-to-month (Figure S1Figure S2). We did not account for day-of-the-week effects in sampling diagnosis as there is enough variance in sampling to obtain CS case date of diagnosis that any effect would be minimal.

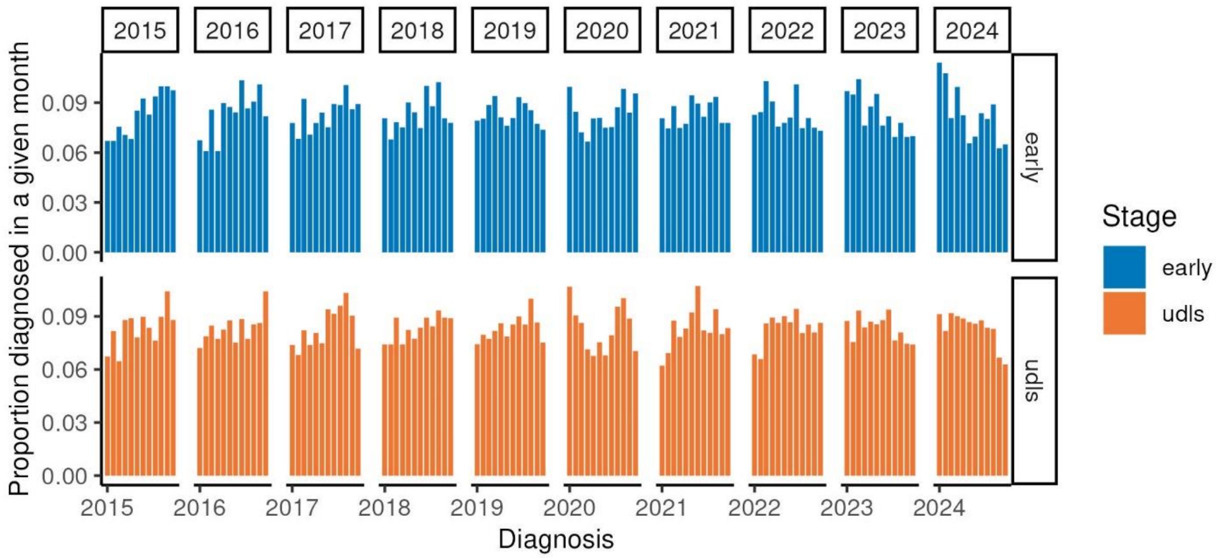


Figure S2. Monthly distribution of diagnosis dates for female acquired syphilis cases between 2015–2024

Treatment for syphilis during pregnancy is classified into four categories in California: (1) no treatment, (2) inadequate treatment (i.e., treatment with incorrect medication, dosage, or spacing between doses),

(3) treatment too late for the prevention of congenital syphilis but adequate treatment for syphilis during pregnancy (i.e., adequate treatment began fewer than 30 days before delivery of the infant), (4) adequate treatment for prevention of congenital syphilis and syphilis during pregnancy. Adequate treatment for syphilis during pregnancy requires the correct medication, dosage, and timing between doses. For early-stage syphilis (i.e., syphilis infection acquired within 12 months; primary, secondary, and early non-primary non-secondary) during pregnancy, adequate treatment requires one dose of penicillin G benzathine 2.4 million units. For late-stage syphilis (i.e., syphilis infection acquired over 12 months ago) or syphilis of unknown duration during pregnancy, adequate treatment requires three doses of penicillin G benzathine 2.4 million units. The three doses must be six to eight days apart. For the prevention of congenital syphilis, maternal treatment must begin 30 or more days before delivery of the infant to be considered adequate.

There are two pathways, maternal and neonatal, for children born from a parent infected with acquired syphilis during pregnancy to be considered CS cases by surveillance criteria. CS cases need to meet the criteria for at least one of two pathways. The neonatal pathway includes (1) demonstration of *Treponema pallidum* by positive darkfield, direct fluorescence antibody (DFA), polymerase chain reaction (PCR), or other special stains, (2) stillbirth after 20 weeks gestational age or fetus weighs greater than 500 grams and birthing parent meets the surveillance case definition and was untreated or inadequate treated, (3) a reactive non-treponemal test (rapid plasma reagin, RPR, or Venereal Disease Research Laboratory, VDRL) and reactive cerebral spinal fluid (CSF)-VDRL, evidence of CS on long bone x-ray, other evidence of CS on physical examination, and/or an elevated CSF white blood cell or protein count, without traumatic tap or other cause, or (4) neonatal/infant/child diagnosis post-discharge with no indication of infection at delivery. The maternal pathway criteria are met if treatment during pregnancy was incomplete (inadequate, late, or no treatment, as described above).

Though CS is largely preventable with adequate and timely maternal treatment, some infants with adequate maternal treatment will still meet neonatal pathway surveillance case criteria and will be considered cases. Conversely, even if the birthing parent's syphilis treatment was incomplete prior to pregnancy, infants may not be considered to have CS if the birthing parent's non-treponemal tests remained non-reactive throughout pregnancy and at delivery. In this situation, the pregnant person does not meet criteria to be considered a syphilis case during pregnancy and the infant does not meet the criteria for congenital syphilis through the neonatal pathway. We determined proportions of pregnancies that resulted in their infant(s) being considered a CS case stratified by treatment outcome of the birthing parent.

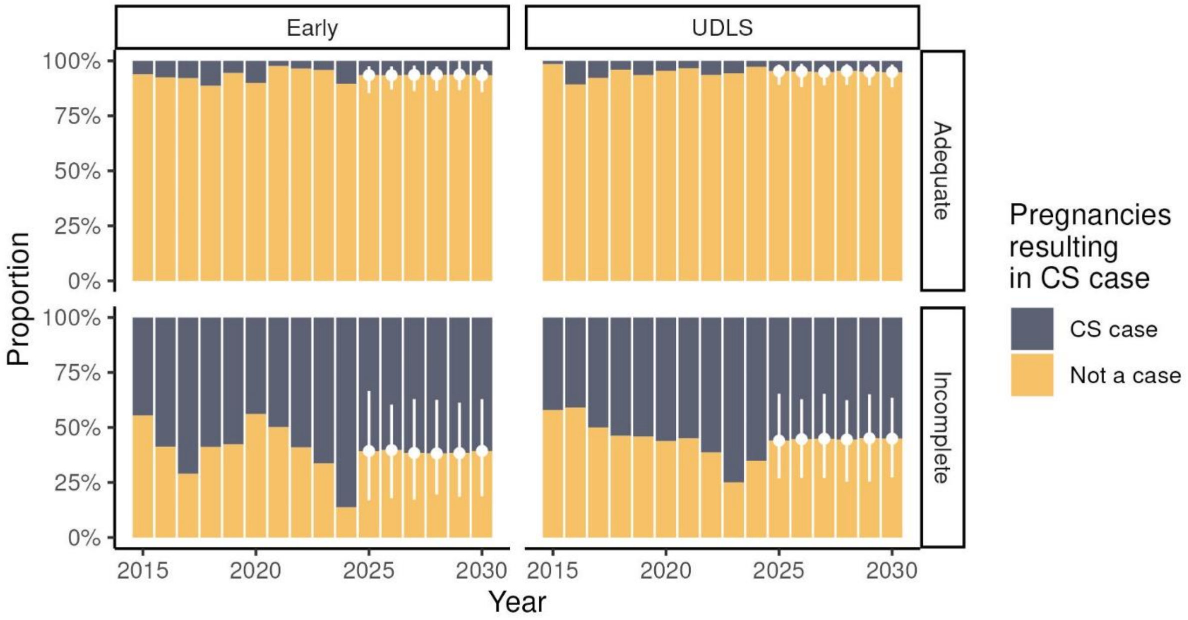


Figure S3. Reported (2015–2024) and projected (2025–2030) proportions of pregnancies resulting in congenital syphilis diagnosis by year, stage of birthing parent infection at diagnosis, and treatment adequacy during pregnancy

Proportions identified as cases despite adequate treatment are similar to estimates from Chicago which found 11.9% of CS cases in 2015–2019 and 9.0% of cases in 2020–2022 had clinical evidence of syphilis despite birthing parent receipt of adequate treatment.[1] The remainder of infants born to adequately treated individuals were categorized as cases averted.

The next stage of sampling is based on 2015–2024 proportions of treatment outcomes for pregnant cases. Direct calculation of adequate treatment during pregnancy was limited to pregnant person records that matched with infant records in our surveillance system. The remaining pregnant cases (32.0%) were missing treatment outcome at birth of their child(ren), so we used the R package *mice* to impute treatment outcome for these individuals. The data were trained on age, year of diagnosis, county of residence, and syphilis stage at diagnosis (early syphilis or UDLS).

For analysis, we simplify the four treatment categories to: 1) adequate treatment and 2) incomplete treatment. Incomplete treatment encompasses no treatment, inadequate treatment, and late treatment. Figure S4 shows the proportion of treatment outcomes for pregnant women with syphilis from surveillance data (2015–2024) and simulated proportions used in proportions of each treatment outcome largely did not change over time for early syphilis, but the UDLS proportion adequately treated reached 50% for the first time in 2023, dropping back down in 2024.

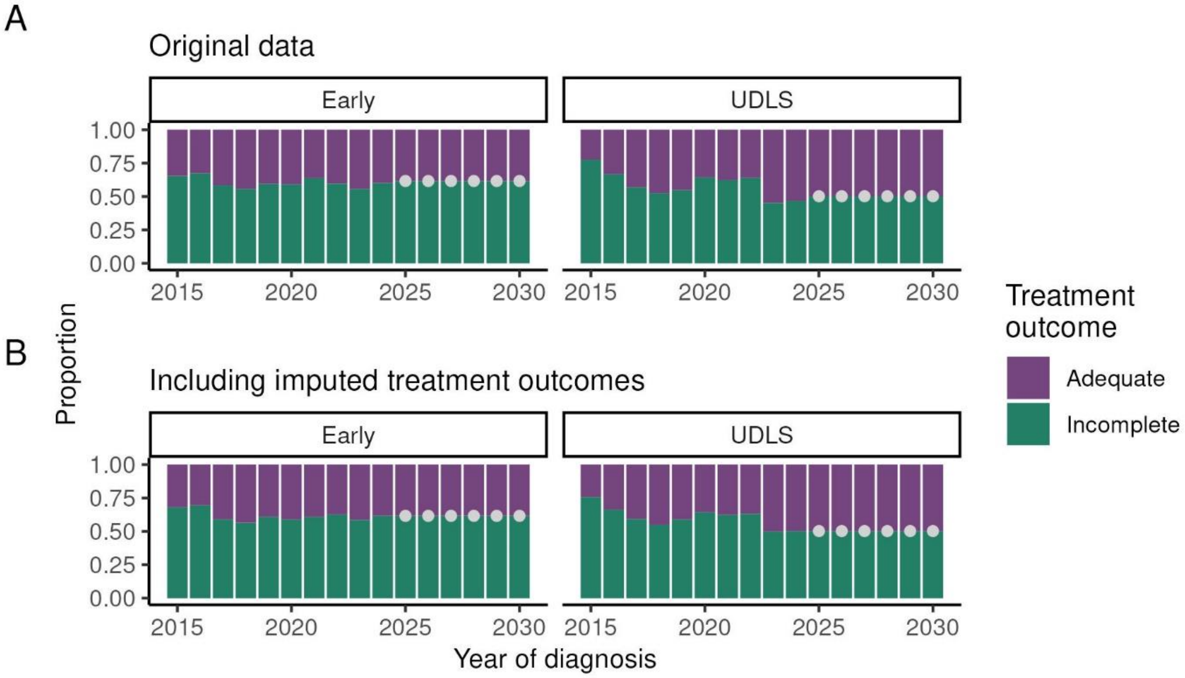


Figure S4. Reported (2015–2024) and projected (2025–2030) proportions of treatment outcomes for pregnant female acquired syphilis cases at date of delivery by stage of infection at diagnosis when considering only reported data (A) or including imputed data (B)

We used the *MCMCprecision* package to fit a Dirichlet distribution to the 2015–2024 data (Figure S5).

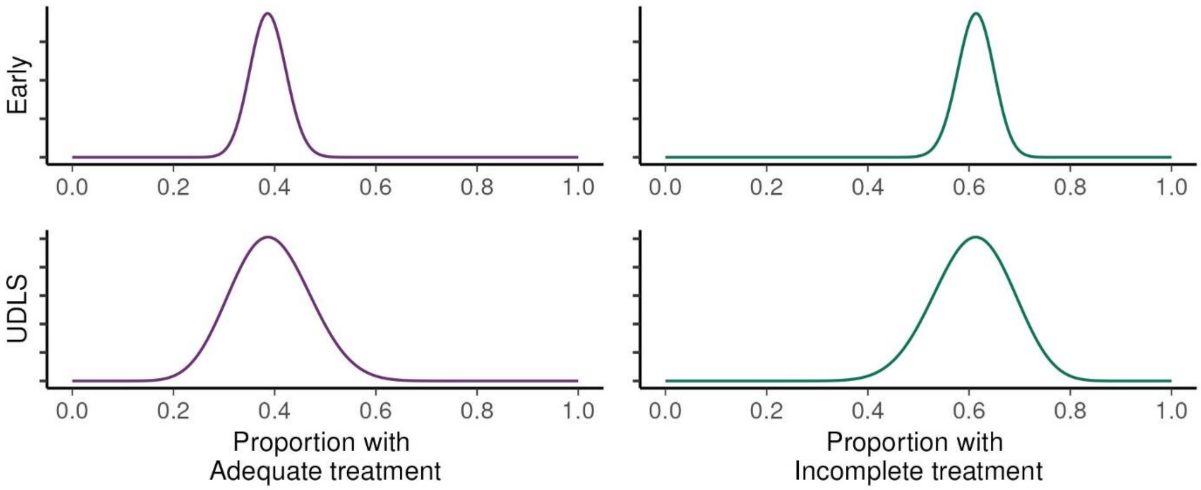


Figure S5. Treatment outcomes among pregnant female syphilis cases

Next, for each of those pregnancies, we determine CS case outcomes. Outcomes are classified as either: live born with no signs of CS, live born with signs of CS, or stillbirth/neonatal death. Cases of late congenital syphilis (onset and diagnosis after two years of age) are not considered in this analysis.

Infants born to pregnant individuals who were projected to have received incomplete treatment prior to the birth of their child are assigned a treatment outcome by sampling from a Dirichlet distribution fit to the 2015–2024 data using the *MCMCprecision* package (Figure S6Figure S6). The modeling framework also includes an option to skip this step and look at all CS cases without stratifying by outcome.

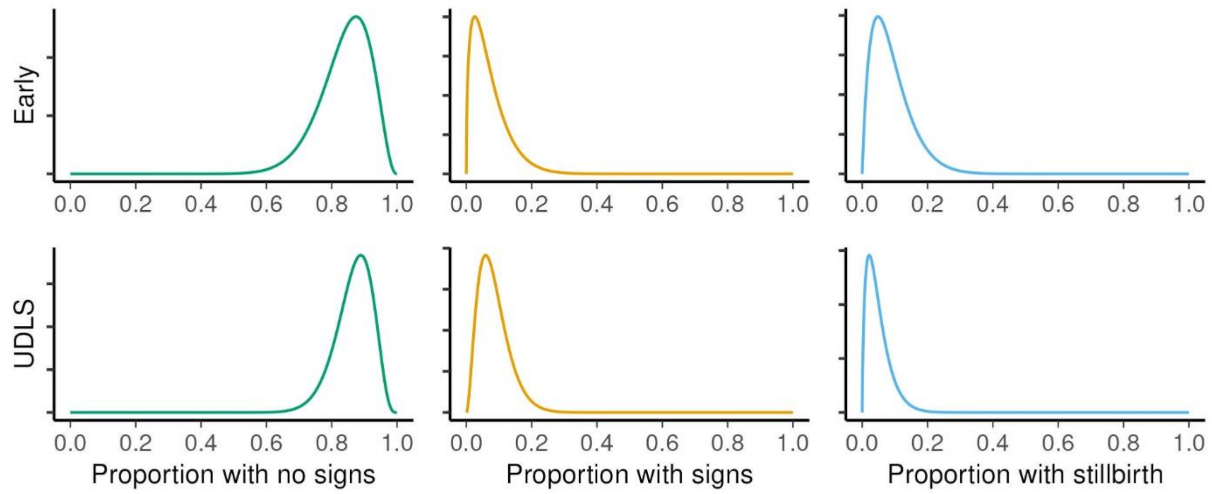


Figure S6. Outcomes for congenital syphilis cases born to a pregnant woman who was not adequately treated for their syphilis infection during pregnancy by birthing parent stage of infection at diagnosis, 2015–2024

Next, we needed to provide an estimate of the timing of when the CS cases would be diagnosed. Most CS cases are diagnosed at birth, so we calculated an estimated date of delivery (EDD) for each pregnant individual based birthing parent's date of diagnosis – days pregnant at diagnosis + length of pregnancy (i.e. 280 days). Days pregnant at diagnosis was sampled from the density distributions shown in **Error! Reference source not found.**. Infection in utero can occur as early as two weeks gestation.

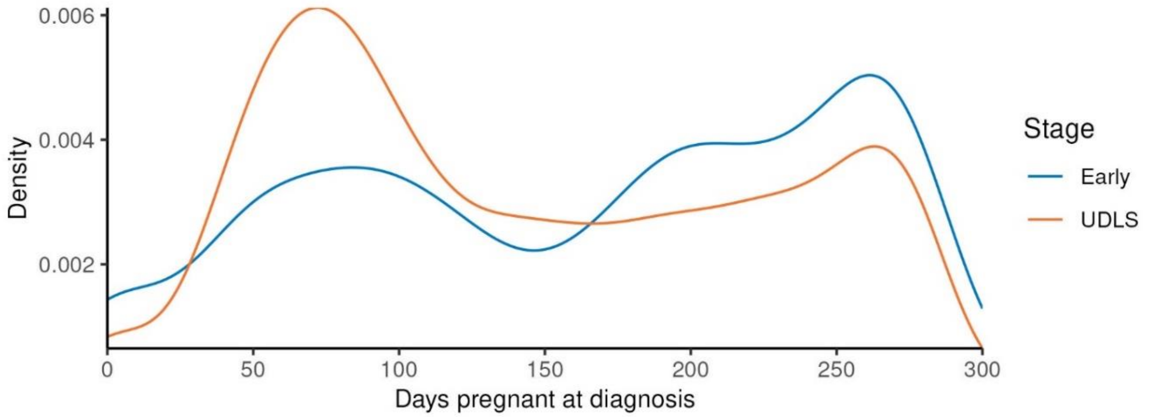


Figure S7. Density of days pregnant at diagnosis among female acquired syphilis cases diagnosed between 2015–2024 by infection stage at diagnosis

We additionally considered that stillbirth/neonatal death cases might be diagnosed earlier than EDD and calculated from the 2015–2024 data the number of days these cases were diagnosed before their EDD. For pregnant individuals diagnosed in the early stage, this averaged 43.6 days (SD: 56.0 days), while for pregnant individuals diagnosed as UDLS, this averaged 68.6 days (SD: 46.0 days). We sampled from a truncated normal distribution with these means and SD to obtain a sampled number of days each stillbirth/neonatal death case would be born early and subtracted that from their estimated EDD to obtain a more realistic timing of diagnosis.

*Calibration to historical data*

To assess how well our model might do to estimate future CS cases given perfect knowledge of the number of female cases and treatment proportions we used data from 2015 through the year prior to the year being analyzed (i.e. 2018 for a 2019 comparison) and visually overlapped those results with the actual CS cases reported for those years. Figure S8A shows the results for this comparison stratifying CS cases by outcome type, while Figure S8B looks at overall total number of cases. When considering just the total number of cases (Figure S8B), the model median was slightly lower than reported CS cases in 2019 and 2020, nearly equivalent in 2021, and slight over the reported CS cases in 2022–2024.

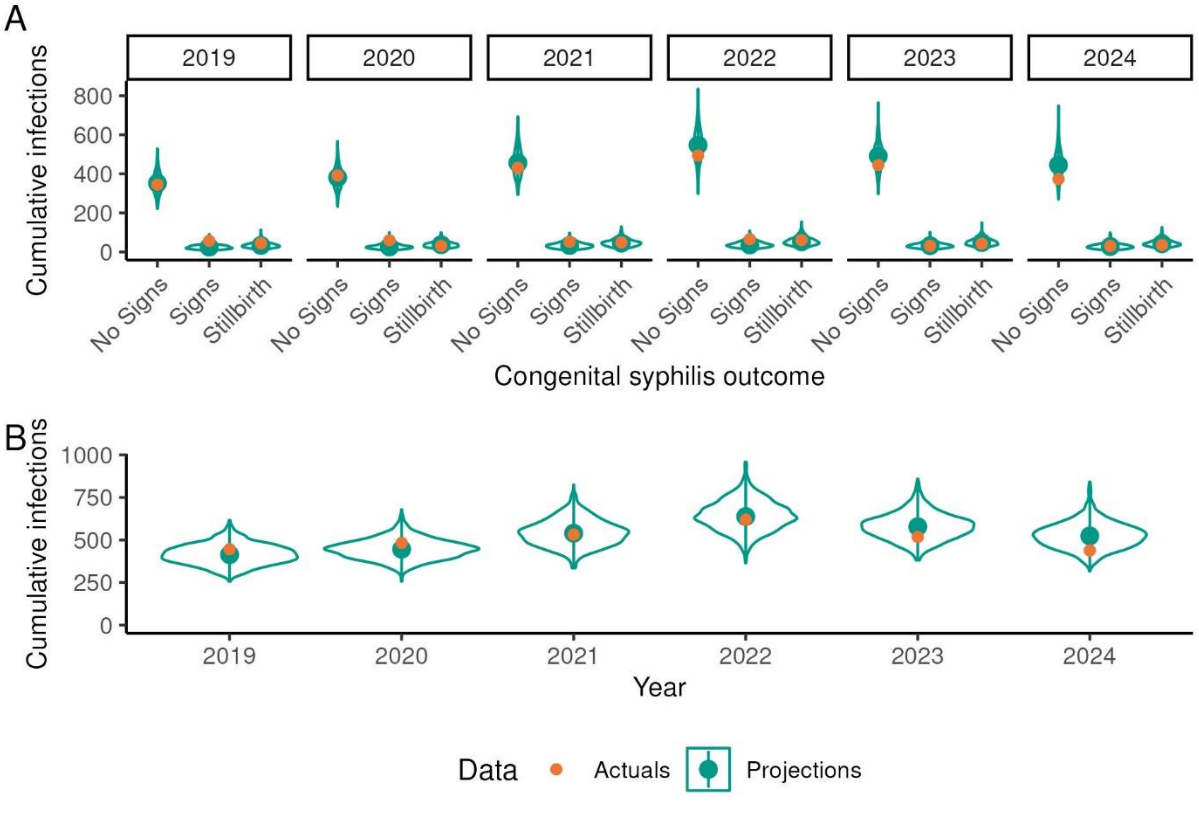


Figure S8. Calibration of congenital syphilis case projections to reported cases 2019–2024 by outcome (A) and for cases overall (B)

We also tracked the projections of female cases in the model (**Error! Reference source not found.**). P rojected case counts closely followed scenario trajectories. With an estimated 50% increase in cases over the six-year projection period (8.3% increase per year), female early syphilis cases reached 2,946 cases and UDLS cases reached 8,890 cases by 2030. With an estimated 50% decrease in cases over the six-year projection period (8.3% decrease per year), female early syphilis cases reached 1,081 cases and UDLS cases reached 3,263 cases by 2030.

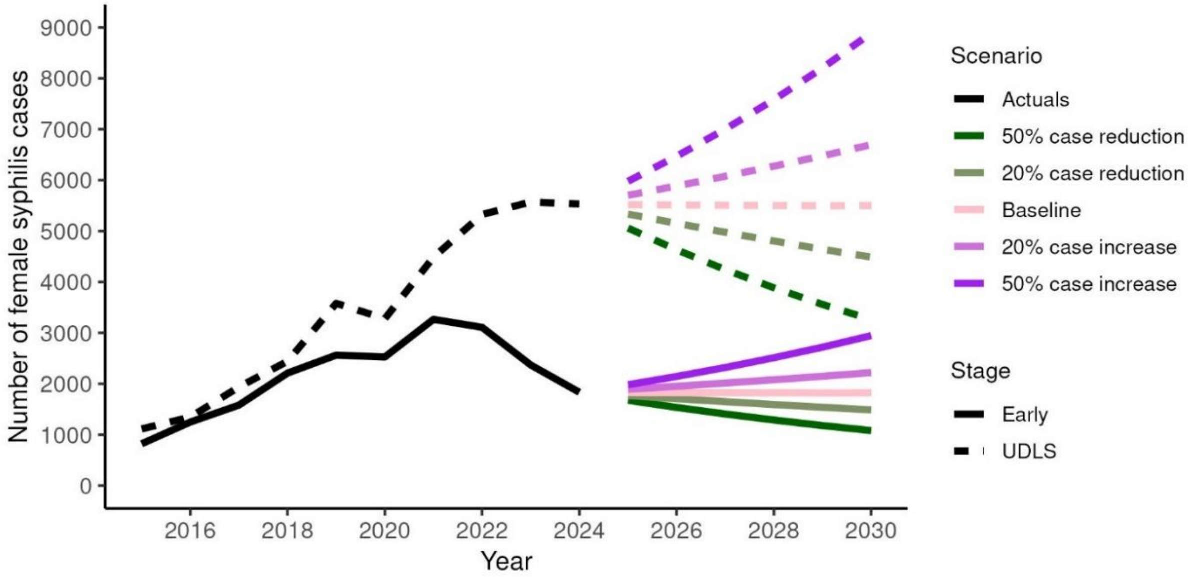


Figure S9. Projected female acquired syphilis cases by stage of infection at diagnosis, 2025–2030

**Hospitalization discharge data**

The California Department of Health Care Access and Information (HCAI) Patient Discharge Dataset (PDD) is mostly representative of hospitals in California but does not include Veteran's Administration hospitals. In analyzing the dataset, we found that there are more hospitalizations with diagnoses of CS than there are reported CS cases, even after attempting to deduplicate hospitalizations to the infant level (Figure S1010). Thus, when using charge data to roughly estimate the potential future hospitalization charges associated with projected CS cases, we felt it was reasonable to assume that on average there would be at least one hospitalization per reported case, particularly given the recent survey of US pediatric physicians that found that 94.1% would choose to administer aqueous penicillin G for 10 days if they believed an infant were highly probable for congenital syphilis.[2]

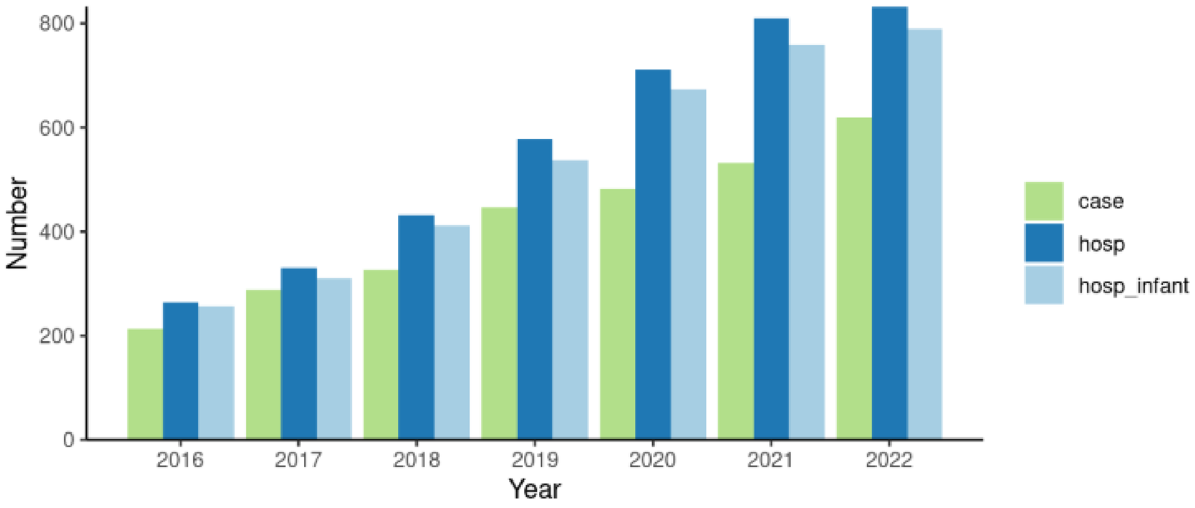


Figure S1010. Hospitalizations, cases, and hospitalizations deduplicated to the infant level in California by year, 2016–2023

In the main article, we present potential future hospitalization charges based on the median charge for any hospitalization with a diagnosis of CS. There is no best practice for obtaining charges related to the recommended 10 days of hospitalization after birth for infants diagnosed with CS to receive treatment, but we found in our analyses that not all admissions occurred within the first day of birth and some were considered transfers where the initial primary diagnosis was the birth itself, and then for a secondary hospitalization the primary diagnosis might be CS or some other diagnosis with CS nonetheless among the other diagnosis codes. Depending on the hospital's testing practices, delays in the receipt of test results for the birthing parent or infant could potentially delay starting treatment.

**Considerations related to diagnosis of CS**

Risk of fetal infection increases with gestational age,[3] and diagnosis of CS is complicated by the presence of birthing parent syphilis antibodies in newborns. Thus, treatment decisions for infants born to a parent who received late, inadequate, or no treatment for syphilis during pregnancy are typically based on a combination of factors, including comparison of birthing parent and neonatal nontreponemal serologic titers at delivery and the presence or absence of clinical, laboratory, and radiographic evidence of syphilis in the neonate.
